# Cardiopulmonary hospitalization risks from wildfire and non-wildfire PM_2.5_ in 20 US states

**DOI:** 10.1101/2025.07.15.25331618

**Authors:** Min Zhang, Edgar Castro, Minghao Qiu, Mahdieh Danesh Yazdi, Boyuan Li, Rosalind J. Wright, Joel D. Schwartz, Robert O. Wright, Yaguang Wei

## Abstract

**Importance:** Given the increasing wildfire activity in the US, assessment of the health impacts of wildfire-specific fine particulate matter (PM_2.5_), a growing source of surface air pollution, and its relative toxicity compared to non-wildfire PM_2.5_ is needed to support mitigation strategies.

**Objective:** To investigate associations of long-term exposure of wildfire-specific and non-wildfire PM_2.5_ with cardiopulmonary hospitalization risks.

**Design, Setting, and Participants:** We obtained over 89 million cardiopulmonary hospitalizations for residents across 20 US states from 2006 to 2019 from the State Inpatient Databases. We assigned estimated 2-year average concentrations of wildfire-specific and non-wildfire PM_2.5_ to each hospitalization based on residential ZIP codes to characterize exposure levels. We used a self-controlled design, which is robust to unmeasured confounding, to assess the associations.

**Exposures:** 2-year moving average exposures to wildfire-specific and non-wildfire PM_2.5_ from the year of hospitalization to the prior year.

**Main Outcomes and Measures:** The hospitalizations for cardiovascular (ischemic heart disease, cerebrovascular disease, heart failure, arrhythmia, other cardiovascular diseases) and pulmonary diseases (acute respiratory infections, pneumonia, chronic obstructive pulmonary disease [COPD], asthma, other respiratory diseases) were identified based on the first 3 diagnosis codes at discharge.

**Results:** Wildfire-specific PM_2.5_ had stronger effects than non-wildfire PM_2.5._ Specifically, each 1-µg/m^3^ increase in 2-year wildfire-specific PM_2.5_ was significantly associated with increased hospitalization risks for all cardiopulmonary diseases, with relative risk ranging from 1.100 (95% CI: 1.091, 1.108) for heart failure to 1.160 (95% CI: 1.142, 1.178) for asthma. In comparison, a 1 µg/m^3^ increase in non-wildfire PM_2.5_ was associated with increased hospitalization risks for all cardiopulmonary diseases, but with relative risks ranging from 1.047 (95% CI: 1.042, 1.051) for COPD to 1.085 (95% CI: 1.082, 1.088) for hypertension. Stronger effects of both wildfire-specific and non-wildfire PM_2.5_ were observed among minorities, individuals with obesity or diabetes, and those living in metropolitan areas, those with fewer years of education, and more deprived communities.

**Conclusions:** Long-term exposure to wildfire-specific PM_2.5_ poses a greater risk of cardiopulmonary hospitalization than PM_2.5_ from non-wildfire sources. Greater effort should be placed on wildfire management, with particular focus on strategies to reduce smoke in addition to traditional air quality control strategies.

**Key point:** *Questions:* Does long-term exposure to fine particulate matter from wildfire and non-wildfire sources affect the risk of cardiopulmonary hospitalization differently?

*Findings:* Based on over 89 million hospitalization records for the residents of 20 US states from 2006 to 2019, exposure to fine particulate matter from wildfire was associated with a greater risk of cardiopulmonary hospitalization, compared to those from non-wildfire sources.

*Meaning:* Fine particulate matter from wildfire sources poses a greater health threat than those from non-wildfire sources; greater effort should be placed on wildfire management in addition to relying solely on traditional air quality control strategies.

## Introduction

Fine particulate matter (PM_2.5_), a major air pollutant, has been identified as the leading environmental risk factor for cardiopulmonary diseases.^1–3^ PM_2.5_ can be drawn deep into the lungs and may pass into the bloodstream, damaging vital organs such as the lungs and heart.^4^ Ambient PM_2.5_ originates from various sources, including traffic, industrial activities, and agriculture. More recently, wildfires are emerging as a significant source of PM_2.5_ due to their increasing frequency and intensity driven by the changing climate,^5–7^ and the expansion of human activity into fire regions.^8^ Globally, the extent of fire-prone areas is expected to expand by 29% towards the late 21st century.^9^ In many parts of the US, wildfire smoke has accounted for over half of the annual PM_2.5_ concentration in extreme smoke years, and has led to reversal of the declining trend in ambient PM_2.5_ over the last two decades due to the Clean Air Act.^10^ As a major toxic component of wildfire smoke, PM_2.5_ can be transported over hundreds of miles, posing a threat to public health beyond the active fire region.^11–13^

Wildfire-specific PM_2.5_ contains higher levels of submicron particles, carbonaceous material (i.e., elemental and organic carbon), and polar organic compounds compared to non-wildfire PM_2.5_.^14–17^ Toxicological studies have shown that, at equivalent doses, wildfire-specific PM_2.5_ induces greater cardiopulmonary toxicity than PM_2.5_ from other sources.^18,19^ However, large-scale evidence comparing the relative toxicity of wildfire-specific and non-wildfire PM_2.5_ is limited.^20,21^ In addition, most existing studies on health effects of wildfire-specific PM_2.5_ have focused on short-term impacts,^13,22,23^ which overlooked the potential accumulation of particles in the body and their sustained effects over time. Addressing these gaps is critical to fully quantify health impacts of wildfires and support the development of long-term strategies to mitigate these impacts in response to the increasing wildfire activity.

In this study, we utilized hospitalization data for the residents of 20 US states between 2006 and 2019 to explore the long-term effects of exposure to wildfire-specific and non-wildfire PM_2.5_ on hospitalization risks of all major types of cardiovascular and pulmonary diseases. We linked hospitalization records with two spatiotemporal models of ambient wildfire-specific and non-wildfire PM_2.5_ levels to estimate each patient’s exposure levels. We used a self-controlled study design that inherently adjusts for time-invariant unmeasured confounders, such as genetics, while also controlling for measured covariates.

## Methods

### Study Population

The hospitalization data of 20 US states (Arizona, California, Colorado, Delaware, Georgia, Iowa, Indiana, Kentucky, Maryland, Michigan, Minnesota, North Carolina, Nebraska, New Jersey, New York, Oregon, Rhode Island, Vermont, Washington, and Wisconsin) from 2006 to 2019 was obtained from the State Inpatient Databases (SID). The SID, a set of state-specific databases developed as part of the Healthcare Cost and Utilization Project (HCUP), contains inpatient discharge records from community hospitals within each participating state.^24^ The SID represents over 95% of all inpatient discharges nationwide.^25^ The availability of SID data varies by year and state, and detailed information is provided in the Supplementary Materials (eTable 1 in **Supplement 1**).

Individual-level elements, including hospital admission year, age, sex, residential zip code, race/ethnicity (White, Black, Hispanic, or other), patient location (metropolitan areas, micropolitan areas, or non-Core Based Statistical Areas [non-CBSA]), diagnosis codes and died during hospitalization (yes vs. no), were extracted from the SID. The cardiopulmonary hospitalizations were identified using the first 3 diagnosis codes at discharge from the *International Classification of Diseases, Ninth Revision or Tenth Revision (ICD-9 or ICD-10)*. The specific ICD codes can be found in eTable2 (**Supplement 1**). The cardiopulmonary diseases of interest included ischemic heart disease, cerebrovascular disease, heart failure, arrhythmia, all other cardiovascular diseases, acute respiratory infections, pneumonia, chronic obstructive pulmonary disease (COPD), asthma, and other respiratory diseases. In addition, co-existing conditions, including obesity or diabetes, were also identified from all secondary diagnosis codes at discharge.

This study was approved by the Institutional Review Board at the Icahn School of Medicine at Mount Sinai. The study followed the Strengthening the Reporting of Observational Studies in Epidemiology (STROBE) reporting guidelines.

### Wildfire-specific PM_2.5_

We obtained daily, ground-level wildfire-specific PM_2.5_ concentrations at a 10 × 10 km^2^ spatial resolution for the contiguous US from 2006–2019 from an open source. These data were estimated using a machine learning model that incorporated ground measurements of PM_2.5_, satellite, and reanalysis data sets^26^. The model achieved reasonable predictive capability, yielding a spatial out-of-sample R^2^ of 0.67 and a within-location R^2^ of 0.65. The grid-level wildfire-specific PM_2.5_ concentrations were aggregated to ZIP Code Tabulation Areas (ZCTAs) using population- and intersection area-weighted averages based on TIGER/Line Shapefiles from the US Census Bureau. These estimates were mapped to zip codes and then linked to the subjects based on their residential address recorded in the SID. For each hospitalization, the exposure window of interest was defined as the 2-year averages of wildfire-specific PM_2.5_ concentrations, covering both the year of hospitalization and the year before.

### Non-Wildfire PM_2.5_

We subtracted the wildfire-specific PM_2.5_ predictions from the all-sourced PM_2.5_ concentrations to obtain the estimated non-wildfire PM_2.5_ concentrations. Those with negative values, which accounted for less than 0.01% of the data, were excluded from this study. We developed high resolution (1 km × 1 km) daily all-sourced PM_2.5_ estimates across the contiguous US during 2006-2019 from a spatio-temporal ensemble model.^27,28^ In brief, a set of predictors from satellite observations, land-use regression, meteorological data, and chemical transport models was used as input for three machine learning algorithms to estimate PM_2.5_ concentrations. The outputs from these models were then integrated using a geographically weighted generalized additive model, which accounts for spatial heterogeneity. The final model demonstrated well performance, with an average cross-validation R^2^ of 0.82. These grid-level estimates were aggregated to the zip code level, and 2-year averages were assigned to each hospitalization.

### Covariates

We extracted neighborhood-level characteristics from databases generated by US Decennial Census for 2000, 2010, and 2020.^29–32^ For the remaining years, values were estimated using linear interpolation. The following zip code-level variables were included in this study: percentage of White, percentage of Black, percentage of the population aged ≥65 years who graduated high school, median household income, and Townsend Deprivation Index (TDI). TDI, an indicator of overall deprivation, is calculated based on four key elements: unemployment, overcrowded households, non-ownership of a car, and non-ownership of a home. A higher score indicates a greater level of deprivation.^33^ These variables were adjusted as potential confounders.

Previous studies have linked ambient temperature to cardiopulmonary outcomes.^24,34,35^ Temperature also directly influences wildfire activity and therefore acts as a confounder. The daily maximum and minimum temperatures (℃) at 1-km^2^ grid cells from 2006 to 2019 were derived from Daymet V4, which was developed by the Oak Ridge National Laboratory.^36^ Daily average temperatures were obtained by averaging the minimum and maximum values. Using daily average temperatures, we calculated the seasonal mean and standard deviation for summer (June, July, and August) and winter (December of the previous year, January, and February) in each year. These meteorological data were aggregated to the zip code level to align with the spatial resolution of hospitalization records.

### Statistical Analysis

The self-controlled design adjusts for all time-invariant and slowly varying confounders through self-matching, regardless of whether these confounders are measured or not. In this study, we extended the traditional case-crossover design, a self-controlled design originally developed for studying short-term effects and widely used, to evaluate long-term effects of 2-year average exposures to wildfire-specific and non-wildfire PM_2.5_ on cardiopulmonary hospitalization risks. Specifically, for each case year (year of hospitalization), the years before and after served as that hospitalization’s own controls. With this self-controlled design, unmeasured time-invariant and slowly varying confounders, such as genetics, chronic coexisting conditions (e.g., diabetes), and smoking status, are inherently adjusted by design. The symmetric bidirectional selection of control years before and after a case year removed the unmeasured confounding from long-term temporal trends (e.g., gradual changes in population-level wildfire smoke level). Selecting one single year before and after the case minimized temporal variations in unmeasured confounders.

We constructed a dataset for each disease and fitted a conditional logistic regression to estimate the simultaneous effects of long-term wildfire-specific and non-wildfire PM_2.5_ on the hospitalization risk, adjusting for measured time-varying confounders, including neighborhood-level covariates, temperatures, and calendar year. Computational details for the fast implementation of conditional logistic regression are provided in supplement 1.

We further conducted stratified analyses based on individual-level factors (age, sex, race/ethnicity, patient location, comorbidity with obesity, and comorbidity with diabetes), disease severity (died/not died during hospitalization) and neighborhood-level characteristics (upper or lower quartiles of percentage of the population who graduated high school and TDI) to determine whether the associations varied across different groups. The differences between subgroups were assessed by the Z-test or the Wald test, as appropriate.

We reported estimated relative risk (RR) of cardiopulmonary hospitalization associated with a 1 μg/m^3^ increase in 2-year average wildfire-specific and non-wildfire PM_2.5_ concentrations. Considering the increased risk of false positives from multiple testing, we applied a Bonferroni correction to adjust 95% confidence intervals (CIs).

## Results

A total of 57,264,966 hospitalizations for cardiovascular diseases and 32,691,394 for pulmonary diseases were included in this study (**Table 1**). The largest proportion of hospitalization for cardiovascular diseases and pulmonary diseases were hypertension (27.5%) and other respiratory diseases (42.4%), respectively. Patients hospitalized for acute respiratory diseases and asthma were younger than those for other diseases. The majority of patients were White and residents of metropolitan areas. Comorbidity with obesity was most prevalent among patients hospitalized for asthma (12.0%), while diabetes was most prevalent among those hospitalized for heart failure (32.8%).

**Table 1.** Characteristics for hospitalized patients.

| Characteristics | Cardiovascular diseases (CVD) |  |  |  |  |  | Respiratory diseases (RD) |  |  |  |  |
| --- | --- | --- | --- | --- | --- | --- | --- | --- | --- | --- | --- |
|  | Ischemic heart disease | Cerebrovascular disease | Heart failure | Arrhythmia | Hypertension | Other cardiovascular diseases | Acute respiratory infections | Pneumonia | COPD | Asthma | Other respiratory diseases |
| Number of individuals, n | 8300356 | 5171077 | 9867248 | 7632375 | 15754362 | 10539548 | 1404055 | 7976711 | 6048248 | 3416258 | 13846122 |
| Age, y, mean (SD) | 68.5 (13.6) | 69.6 (15.2) | 72.8 (14.6) | 71.4 (15.8) | 65.2 (15.5) | 64.9 (18.0) | 26.3 (31.5) | 64.5 (23.2) | 68.8 (12.7) | 40.9 (25.2) | 63.8 (21.5) |
| Sex, n |  |  |  |  |  |  |  |  |  |  |  |
| Female | 3278182 | 2650863 | 5004769 | 3660338 | 8229910 | 5225091 | 706010 | 4108802 | 3268094 | 2200754 | 7105455 |
| Male | 5021667 | 2519843 | 4861715 | 3971418 | 7522757 | 5313291 | 697943 | 3867166 | 2779638 | 1215106 | 6739172 |
| Missing | 507 | 371 | 764 | 619 | 1695 | 1166 | 102 | 743 | 516 | 398 | 1495 |
| Race/ethnicity, n |  |  |  |  |  |  |  |  |  |  |  |
| White | 5726781 | 3318759 | 6505047 | 5559589 | 9465162 | 6880790 | 692307 | 5403834 | 4525708 | 1620777 | 9414885 |
| Black | 848768 | 799576 | 1580404 | 697758 | 3024803 | 1578797 | 226230 | 958272 | 604069 | 803557 | 1693453 |
| Hispanic | 457647 | 296785 | 503589 | 325244 | 1154959 | 606597 | 211843 | 501809 | 192384 | 442716 | 833581 |
| Other | 499943 | 303494 | 448917 | 330025 | 938269 | 524991 | 121507 | 412588 | 189628 | 242964 | 732452 |
| Missing | 767217 | 452463 | 829291 | 719759 | 1171169 | 948373 | 152168 | 700208 | 536459 | 306244 | 1171751 |
| Patient location, n |  |  |  |  |  |  |  |  |  |  |  |
| Metropolitan areas | 6612443 | 4269116 | 7998579 | 6223616 | 13109443 | 8761543 | 1171856 | 6203349 | 4548566 | 2980905 | 11345511 |
| Micropolitan areas | 973852 | 530530 | 1086703 | 819194 | 1554821 | 1051594 | 132460 | 987304 | 814618 | 265354 | 1495092 |
| Non-CBSA | 710461 | 369970 | 779060 | 586538 | 1085088 | 723156 | 99417 | 784077 | 682621 | 168903 | 1002881 |
| Missing | 3600 | 1461 | 2906 | 3027 | 5010 | 3255 | 322 | 1981 | 2443 | 1096 | 2638 |
| Died during hospitalization, n | 293836 | 258276 | 287764 | 179291 | 112885 | 273830 | 1423 | 409586 | 89868 | 8201 | 1317231 |
| Comorbidity with obesity, n | 774415 | 306140 | 939909 | 655384 | 1740327 | 953261 | 66801 | 474999 | 569813 | 410053 | 1078374 |
| Comorbidity with diabetes, n | 2560949 | 1422334 | 3233121 | 1722448 | 4323743 | 2337610 | 130744 | 1671658 | 1478897 | 470441 | 2622252 |
| Neighborhood-level statistics, mean (SD) |  |  |  |  |  |  |  |  |  |  |  |
| Percentage of White, % | 71.2 (24.5) | 68.3 (25.6) | 68.3 (26.0) | 72.5 (23.3) | 65.5 (27.0) | 68.8 (25.3) | 63.9 (27.2) | 70.4 (24.8) | 72.9 (23.8) | 60.8 (28.7) | 69.2 (24.4) |
| Percentage of Black, % | 12.7 (19.1) | 14.7 (21.0) | 15.3 (21.7) | 11.8 (18.1) | 16.7 (22.5) | 14.4 (20.8) | 16.1 (21.5) | 13.2 (19.4) | 13.0 (19.1) | 19.8 (24.2) | 13.2 (19.3) |
| Percentage of the population who graduated high school, % | 85.6 (8.4) | 85.9 (8.3) | 85.5 (8.4) | 86.7 (8.05) | 85.2 (8.7) | 86.1 (8.3) | 83.8 (9.7) | 85.2 (8.5) | 84.8 (8.3) | 83.5 (9.6) | 86.0 (8.3) |
| Median household income, \$ | 56726.8<br>(23379.9) | 57187.2<br>(23627.4) | 55516.5<br>(22987.9) | 58990.5<br>(24311.3) | 56687.8<br>(23728.4) | 57508.0<br>(23889.9) | 54716.8<br>(23244.8) | 54949.8<br>(22452.5) | 52519.0<br>(20819.6) | 53429.9<br>(22936.5) | 57081.7<br>(23316.3) |
| Townsend deprivation index | 1.25 (3.15) | 1.32 (3.12) | 1.47 (3.25) | 1.04 (2.98) | 1.68 (3.52) | 1.31 (3.16) | 2.03 (3.76) | 1.28 (3.05) | 1.15 (2.80) | 2.63 (4.24) | 1.18 (2.88) |
| Environmental data, median (ranges) |  |  |  |  |  |  |  |  |  |  |  |
| Wildfire-specific PM<sub>2.5</sub>, µg/m<sup>3</sup> | 0.30 (0.00, 11.15) | 0.29 (0.00, 11.15) | 0.31 (0.00, 11.15) | 0.30 (0.00, 11.15) | 0.29 (0.00, 11.15) | 0.30 (0.00, 11.15) | 0.29 (0.00, 10.26) | 0.30 (0.00, 11.15) | 0.30 (0.00, 11.15) | 0.29 (0.00, 10.26) | 0.29 (0.00, 11.15) |
| Non-wildfire PM<sub>2.5</sub>, µg/m<sup>3</sup> | 8.06 (0.02, 22.78) | 7.96 (0.02, 20.06) | 8.04 (0.02, 20.85) | 7.94 (0.02, 20.67) | 7.99 (0.005, 21.30) | 7.95 (0.02, 20.67) | 8.05 (0.006, 19.24) | 7.96 (0.02, 20.85) | 8.03 (0.005, 20.85) | 8.32 (0.005, 20.85) | 7.78 (0.005, 20.85) |
| Mean summer temperature, °C | 22.89 (11.80, 35.53) | 22.97 (11.44, 35.53) | 22.93 (11.44, 35.53) | 22.82 (11.44, 35.53) | 23.14 (11.47, 35.53) | 22.96 (11.44, 35.53) | 23.07 (11.60, 35.53) | 23.01 (11.44, 35.53) | 22.90 (11.80, 35.53) | 23.11 (11.60, 35.53) | 22.96 (11.44, 35.53) |
| SD of summer temperature, °C | 1.51 (0.001, 4.41) | 1.49 (0.001, 4.35) | 1.51 (0.001, 4.41) | 1.50 (0.001, 4.46) | 1.51 (0.001, 4.46) | 1.50 (0.001, 4.33) | 1.55 (0.001, 4.35) | 1.47 (0.001, 4.46) | 1.44 (0.001, 4.46) | 1.58 (0.001, 4.35) | 1.47 (0.001, 4.41) |
| Mean winter temperature, °C | 1.46 (-19.26, 23.71) | 1.84 (-19.26, 23.23) | 1.49 (-19.22, 23.71) | 1.46 (-19.26, 23.28) | 1.84 (-19.26, 23.71) | 1.63 (-19.26, 23.71) | 1.69 (-19.22, 23.19) | 1.69 (-19.26, 23.22) | 1.43 (-19.26, 23.22) | 1.45 (-19.26, 23.71) | 1.98 (-19.26, 23.71) |
| SD of summer temperature, °C | 2.07 (0.005, 9.44) | 2.08 (0.005, 9.50) | 2.21 (0.005, 9.46) | 2.13 (0.005, 9.60) | 2.13 (0.005, 9.64) | 2.13 (0.005, 9.61) | 2.05 (0.005, 9.37) | 2.13 (0.005, 9.44) | 2.22 (0.005, 9.54) | 2.06 (0.005, 9.42) | 2.14 (0.005, 9.60) |

Among the 20 US states studied, California recorded the highest concentrations of both wildfire-specific PM_2.5_ and non-wildfire PM_2.5_ (**Figure 1**). During the study period, the annual mean wildfire-specific PM_2.5_ was 0.29 (range: 0.00-11.15) μg/m^3^, and 7.78 (range: 0.005-22.78) μg/m^3^ for non-wildfire PM_2.5_ (**Table 1**). The highest temperature in summer was 35.53 ℃, while the lowest temperature in winter is -19.26 ℃.

**Figure 1.**
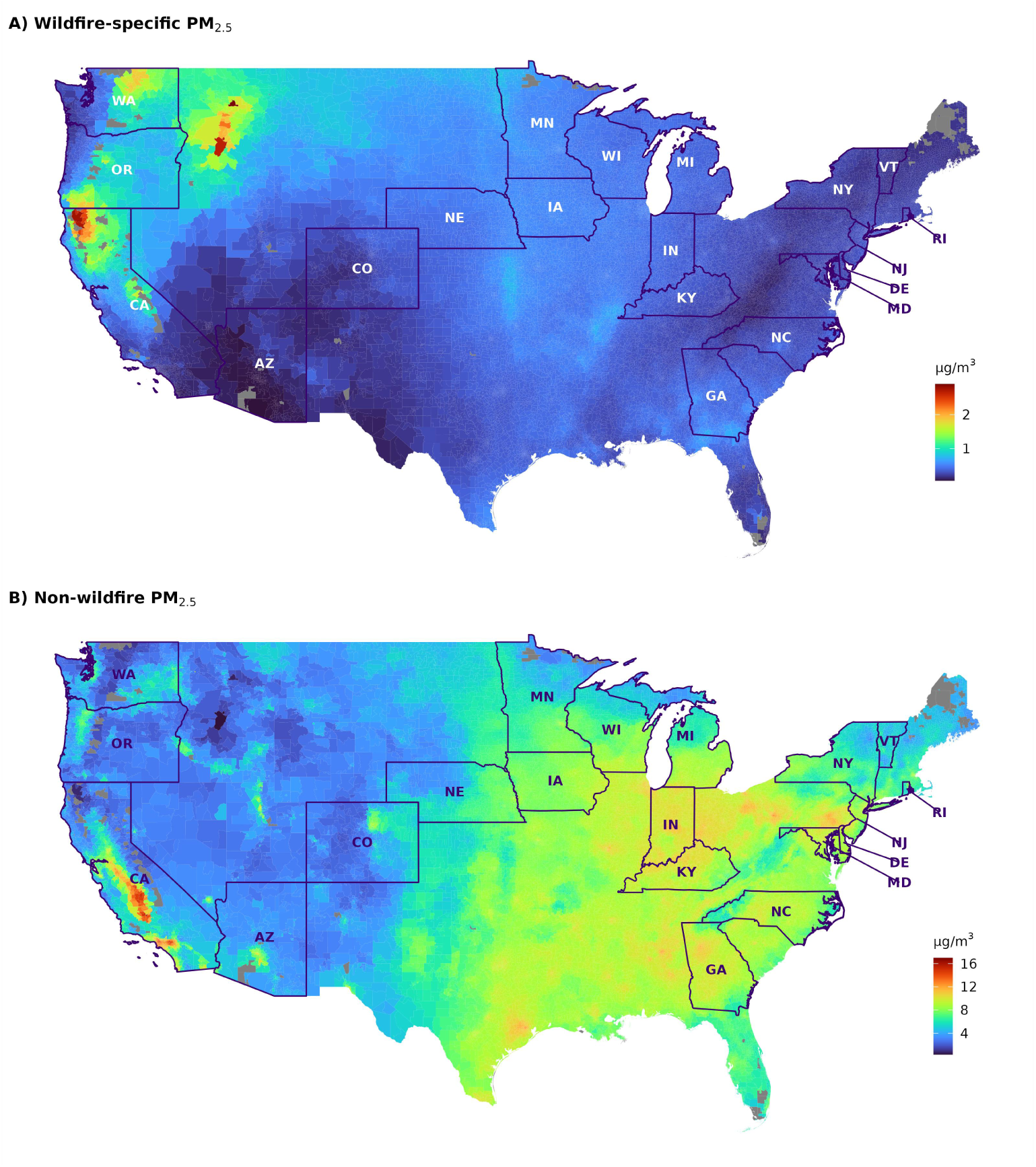
Average concentrations of wildfire-specific PM_2.5_ and non-wildfire PM_2.5_ (μg/m^3^) in the contiguous United States from 2006-2019. The states marked with abbreviations in the box are the ones included in our study.

Compared to non-wildfire PM_2.5_, wildfire-specific PM_2.5_ was associated with greater risks of hospitalizations for all the examined cardiopulmonary diseases (**Figure 2**). Each 1-μg/m^3^ increase in 2-year average wildfire-specific PM_2.5_ concentrations was significantly associated with increased hospitalization risks for cerebrovascular disease (RR, 1.131; 95% CI, 1.119–1.143), ischemic heart disease (RR, 1.129; 95% CI, 1.119–1.139), hypertension (RR, 1.122; 95% CI, 1.115–1.129), arrhythmia (RR, 1.117; 95% CI, 1.108–1.127), other cardiovascular diseases (RR, 1.117; 95% CI, 1.109–1.126), and heart failure (RR, 1.100; 95% CI, 1.091–1.108). For pulmonary diseases, wildfire-specific PM_2.5_ had the greatest impact on asthma, with an RR of 1.160 (95% CI, 1.142–1.178), and a smaller effect on pneumonia, with an RR of 1.109 (95% CI, 1.099–1.118).

**Figure 2.**
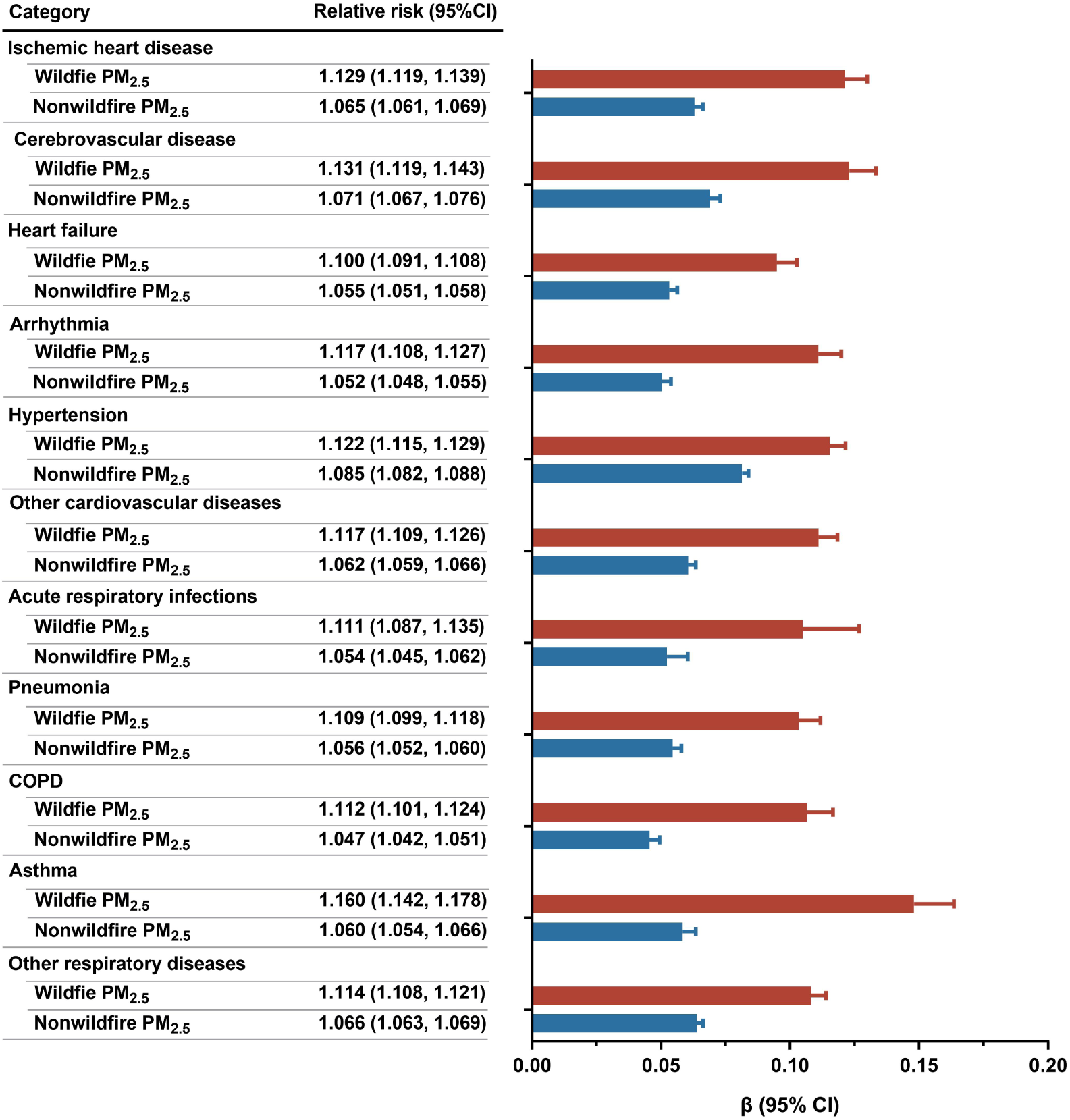
The hospitalization risks of cardiopulmonary diseases associated with a 1-μg/m^3^ increase in 2-year average of wildfire-specific PM_2.5_ and non-wildfire PM_2.5_ concentrations. The model was adjusted for calendar year, percentage of White, percentage of Black, percentage of the population who graduated high school, median household income, townsend deprivation index, mean temperature of summer and winter, and standard deviation of summer and winter temperature.

Positive associations were also observed between non-wildfire PM_2.5_ and hospitalization risks of all cardiopulmonary diseases (**Figure 2**). A 1-μg/m^3^ increase in 2-year average non-wildfire PM_2.5_ concentrations was significantly associated with increased risks of cardiovascular hospitalization: 1.085 (95% CI, 1.082–1.088) for hypertension, 1.071% (95% CI, 1.067–1.076) for cerebrovascular disease, 1.065 (95% CI, 1.061–1.069) for ischemic heart disease, 1.062 (95% CI, 1.059–1.066) for other cardiovascular diseases, 1.055 (95% CI, 1.051–1.058) for heart failure, and 1.052 (95% CI, 1.048–1.055) for arrhythmia. Among all pulmonary diseases, the association was the largest for other respiratory diseases (RR, 1.066; 95% CI, 1.063–1.069), followed by asthma (RR, 1.060; 95% CI, 1.054–1.066), and pneumonia (RR, 1.0576; 95% CI, 1.052–1.060). These results remained robust after changing the degrees of freedom for temperature up to 9, adjusting for annual precipitation and vapor pressure, or identifying hospitalizations based on the principal diagnosis code at discharge (eTable 3-5 in **Supplement 1**).

Hispanic individuals and those from other racial/ethnic groups experienced greater cardiopulmonary hospitalization risks (**Figure 3**). Stronger particle associated risk was observed among people who lived in metropolitan areas (**Figure 3**), communities with lower education attainment, or communities with greater deprivation level (**Figure 4**). Moreover, the presence of co-existing obesity or diabetes increased particle-related hospitalization risks for most types of cardiopulmonary diseases (**Figure 4**). There were few differences in PM_2.5_-related cardiopulmonary hospitalization risks across subpopulations defined by age, sex, and disease severity (**Figure 3**-**4**; eTable 6-14 in **Supplement 1**).

**Figure 3.**
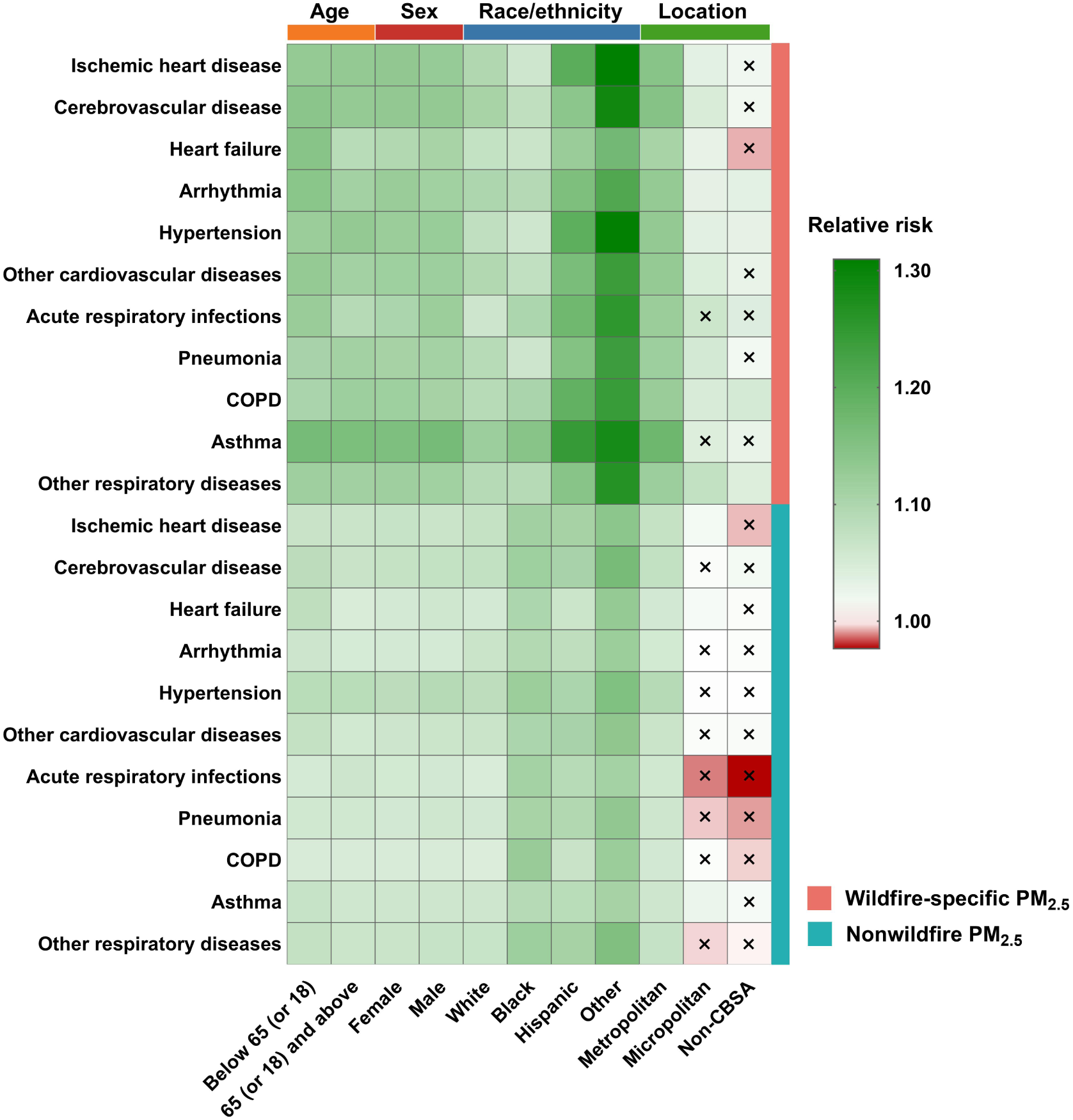
Heatmap of relative risks from subgroup analyses by age, sex, race/ethnicity, and patient location. Different colors in the heatmap represent different values of relative risks. Values equal to 1 are shown in white, and the deeper the color, the further the value is from 1. Boxes marked with a cross symbol (×) denote that the 95% confidence interval contains 1. Age was stratified at 18 years for acute respiratory infections and asthma, and at 65 years for all other disease outcomes. All numerical results, as well as the results of between-group comparisons, can be found in Supplementary eTables 5-8. The Z-test was used when comparing two subgroups, and the Wald test was applied when comparing more than two subgroups.

**Figure 4.**
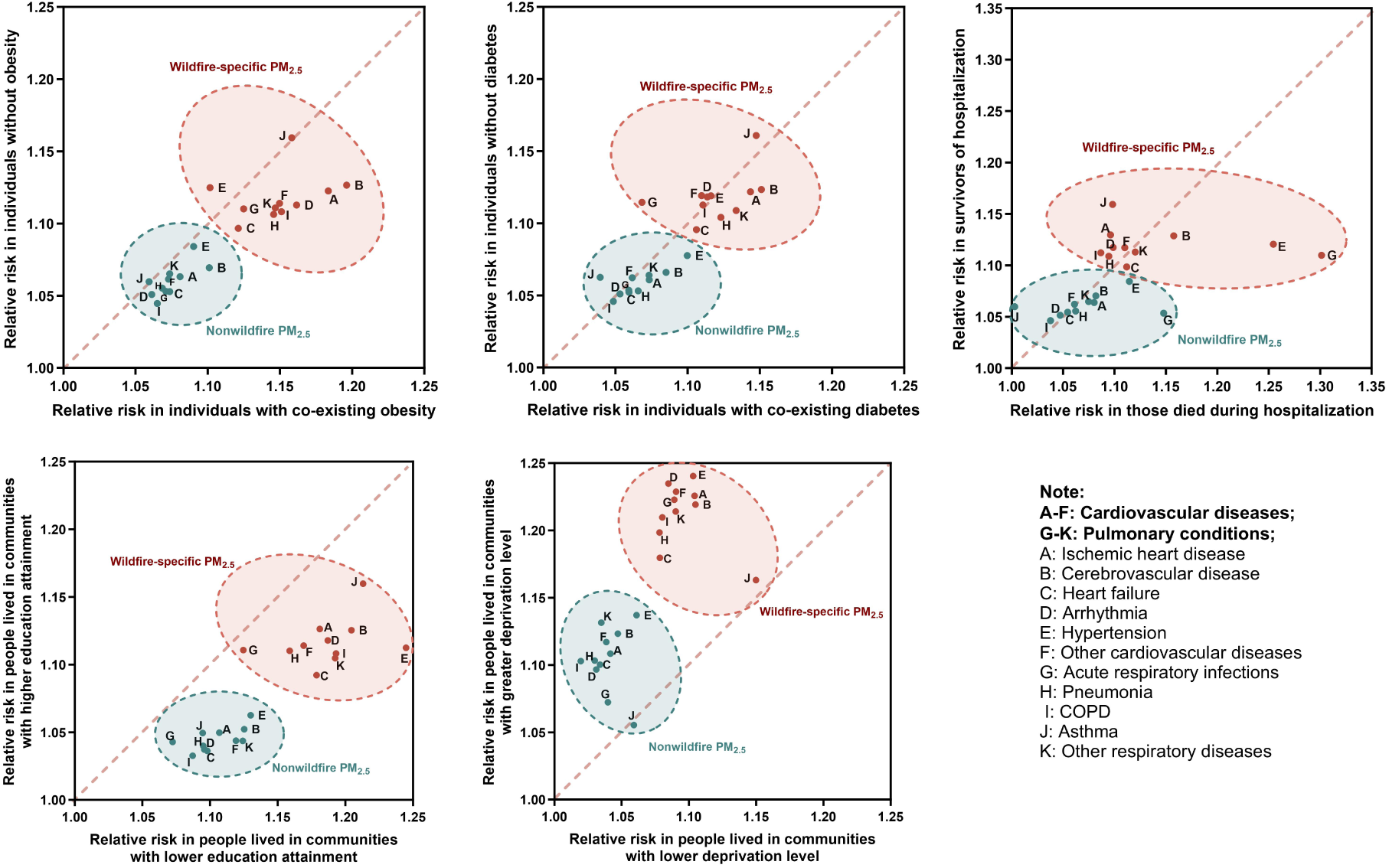
Scatter plot of relative risks from subgroup analyses based on comorbidities, disease severity, and community-level factors. The greater the distance of a point from the diagonal line, the higher the relative risk it represents within a specific subgroup. All relative risks shown have 95% confidence intervals entirely above 1. The percentage of the population who graduated high school was divided into two groups based on the lower quartile (25th percentile): lower and higher educational attainment. Townsend deprivation index was categorized into two groups based on the upper quartile (75th percentile): lower and higher deprivation levels. Detailed numerical results, as well as the results of between-group comparisons, are provided in Supplementary eTables 9–13. The Z-test was used when comparing two subgroups.

## Discussion

In this study across 20 US states, a 2-year average exposure to both wildfire-specific PM_2.5_ and non-wildfire PM_2.5_ was associated with increased hospitalization risks for all major types of cardiopulmonary diseases. Although most wildfires are abrupt events, our findings suggest that their health effects are persistent, and there is a need for long-term wildfire management strategies and protective measures to reduce exposure to smoke. Importantly, we found that wildfire-specific PM_2.5_ posed greater hospitalization risks per unit exposure, suggesting greater effort should be placed on wildfire management in addition to relying solely on traditional air quality control strategies. Currently, these strategies largely exempt wildfire smoke from determining attainment of the National Ambient Air Quality Standards (NAAQS).^37^ Moreover, traditional wildfire management approaches, such as controlled burns, have primarily focused on protecting property rather than minimizing population exposure to smoke–an aspect that should be more fully integrated in future efforts. Greater susceptibility to both wildfire-specific PM_2.5_ and non-wildfire PM_2.5_ was found among minorities, those with co-existing obesity or diabetes, individuals in metropolitan areas, and communities with lower education attainment and greater deprivation level.

We used a self-controlled design to evaluate the long-term effects of exposure. Unlike the conventional case-crossover design, which is primarily used to assess short-term effects,^38^ our design selects control years both before and after the case year to carry out the assessment over a longer time horizon. To account for potential confounding introduced by the extended time frame, we adjusted for a wide range of time-varying confounders, such as calendar year, seasonal temperature, and neighborhood-level statistics. This approach preserves the strengths of the case-crossover design, in particular its ability to control for time-invariant individual-level confounders, while expanding its applicability to study long-term effects.

Our findings are consistent with the emerging evidence on the associations between exposure to wildfire-specific PM_2.5_ and increased risks of pulmonary diseases, especially asthma.^21,39,40^ We observed that a 1-μg/m^3^ increase in wildfire-specific PM_2.5_ concentrations was associated with a 10.9%–16.0% increase in pulmonary hospitalization risks, a magnitude generally greater than that reported in previous studies. A study of eight countries and territories found a 0.25% (RR= 1.0025, 95% CI: 1.0023, 1.0028) increase in all-cause pulmonary hospitalizations per 1-μg/m^3^ increase in the 2-day average concentration of wildfire smoke PM_2.5_.^21^ Another study of 16 US states reported a 2.1%–2.3% increase in hospitalizations for COPD, asthma, and other respiratory diseases associated with a 1-μg/m^3^ increase in 3-month moving average exposure to wildfire smoke PM_2.5_.^41^ The larger magnitudes of associations might be due to the longer exposure window than previous studies. Our earlier work showed that chronic exposure to ambient air toxicants posed substantially larger risks than acute exposures over a few days, potentially due to the accumulation of toxicants in the body and their sustained adverse effects on various organs over time.^42^

For the effects of wildfire emissions on cardiovascular diseases, existing evidence remains less conclusive.^43–46^ Wei et al. found significant associations between 3-month average exposure to wildfire-specific PM_2.5_ and increased hospitalization risks of cerebrovascular disease, hypertension, and other cardiovascular diseases in US.^41^ Alexeeff et al. observed that exposure to elevated PM_2.5_ levels during the Mendocino Complex wildfire was associated with increased cardiovascular hospitalization risks, whereas no significant association was observed during the Camp Fire. Several studies have found little evidence of increased risk during wildfire episodes^47–51^ or in association with short-term exposure to wildfire-specific PM_2.5_.^39,40,52–54^ Previous studies used short exposure windows (typically within one week), which may fail to capture the long-term health impacts of smoke on cardiovascular outcomes. One study suggested that the increase in emergency department visits for cardiovascular diseases associated with high levels of wildfire-related PM_2.5_ exposure appeared on lag day 10, considerably later than the effect observed for pulmonary diseases, which began on lag day 0.^55^ Differences in exposure assessment, study population, and study design may explain varied results across studies.

Our research demonstrated that wildfire-specific PM_2.5_ was associated with a greater risk of cardiopulmonary hospitalizations compared to non-wildfire PM_2.5_. Similar results were reported in a multi-city study across eight countries and territories^21^ and in another US study from Southern California,^20^ which suggested that wildfire-specific PM_2.5_ led to 3 to 10 times higher risk of pulmonary hospital admissions compared to PM_2.5_ from non-wildfire sources. Aligned with these findings, a mouse model study showed that exposure to wildfire-derived PM at doses equivalent to ambient PM induced stronger pulmonary inflammation and histological lung damage.^19^ In mice exposed to PM from smoldering peat fire, lung inflammation associated with endotoxins and reactive oxygen species, reduced cardiac function, and post-ischemia-associated myocardial infarction were observed.^18^ This elevated risk associated with wildfire-specific PM_2.5_ may be attributed to its unique chemical composition, which tends to contain a higher proportion of submicron and ultrafine particles, as well as elevated concentrations of oxidative and pro-inflammatory substances, such as polycyclic aromatic hydrocarbons.^14–17,23,56–58^ Because components of wildfire-specific PM_2.5_ are smaller in size, they may more easily penetrate the human body, which facilitate their adverse effects.

We found that vulnerable subgroups were similar for wildfire and non-wildfire PM_2.5_. Greater susceptibility to both wildfire-specific and non-wildfire PM_2.5_ observed among minorities, residents of communities with lower education attainment, and communities with greater deprivation level can be attributed to structural racism and socioeconomic disparities. Compared to those identifying as white or persons with higher social class, both minorities and persons with lower social class face multiple inequities, including poorer healthcare access, higher pollution burden, fewer neighborhood resources, and fewer resources to mitigate the health impacts of PM_2.5_.^59–61^ In addition, individuals residing in metropolitan areas were more susceptible than those residing in micropolitan areas or non-CBSA. This may be explained by higher levels of social stress in metropolitan areas, which can negatively impact immune responses^62,63^ and thereby increase vulnerability to environmental stressors. Moreover, both wildfire-specific and non-wildfire PM_2.5_ had stronger effects among individuals with co-existing obesity or diabetes, highlighting the synergistic effect of PM_2.5_ exposure and cardiopulmonary risk factors.^64–66^ Notably, individuals with obesity were generally more sensitive to PM_2.5_ exposure than those with diabetes. One possible explanation is that individuals with obesity often have reduced cardiopulmonary function, along with systemic chronic inflammation and elevated oxidative stress, all of which may exacerbate the burden of PM_2.5_ exposure.^66^ Our results also support the finding that both wildfire-specific and non-wildfire PM_2.5_ were associated with deaths caused by hospitalization for cardiopulmonary diseases, although the difference in risk between patients who died during hospitalization and those who survived was minimal.

Our study has several strengths. First, we included a large sample covering residents of all ages from 20 US states, including those most heavily affected by wildfires, such as California, Oregon, and Washington. Additional strength include comprehensive coverage of all major cardiopulmonary conditions and the use of a novel self-controlled design that minimizes bias from unmeasured confounders. Our study also has a few limitations. First, the spatial resolution and predictive performance of our wildfire-specific PM_2.5_ prediction model were suboptimal, which may introduce exposure measurement error. This error for each individual within a zip code should mainly be Berksonian, which may not bias our estimates but may widen the confidence intervals. Second, the most recent data used in this study are nearly six years old, and since the climate has been changing rapidly and wildfires have been worsening, it is unclear whether the effects of wildfires would be the same using more current data.

## Conclusions

This study demonstrates that long-term exposure to wildfire-specific PM_2.5_ poses a greater risk of cardiopulmonary hospitalization than non-wildfire PM_2.5_. Furthermore, both wildfire-specific and non-wildfire PM_2.5_ had stronger effects among minorities, those with co-existing obesity or diabetes, and residents of metropolitan areas, communities with lower lower education attainment, and greater deprivation level. Considering the reversal of air quality improvements driven by rapidly increasing wildfire activity and the greater toxicity of wildfire-specific PM_2.5_ compared to non-wildfire PM_2.5_, greater efforts should be placed on wildfire management in addition to relying solely on traditional air quality control strategies.

## Data Availability

The SID are available upon request to the HCUP. The ZIP code-level wildfire-specific PM2.5 and non-wildfire PM2.5 data are available from the corresponding author on reasonable request. The originally predicted, grid level PM2.5 from all-sourced data are available at https://zenodo.org/records/7884640. Meteorological data, including temperature, precipitation, and vapor pressure, are available at https://daac.ornl.gov/DAYMET/guides/Daymet_Daily_V4.html. Other covariate data are publicly available with sources described in the manuscript.

## Article Information

### Author Contributions

Zhang, Danesh Yazdi, Schwartz, and Wei had full access to all of the data in the study and takes responsibility for the integrity of the data and the accuracy of the data analysis.

*Concept and design:* Zhang, Wei.

*Acquisition, analysis, or interpretation of data:* Zhang, Wei, Castro, Qiu.

*Drafting of the manuscript:* Zhang.

*Critical revision of the manuscript for important intellectual content:* All authors.

*Statistical analysis:* Zhang.

*Obtained funding:* Schwartz, Wright, Wei.

*Administrative, technical, or material support:* Wei, Schwartz, Castro.

*Supervision:* Wei.

### Disclaimer

The authors take sole responsibility for all data analyses, interpretations, and views expressed in this work.

### Data Sharing Statement

see Supplement 2.

### Additional Contributions

We thank the following individuals for their suggestions on drawing heatmap: Jie Yin, Yue-Yan Zhu, Jia-Xing Wang, and Hao Hu. These individuals were not compensated for their role in the study.

