## Supplement for "Cardiopulmonary hospitalization risks from wildfire and non-wildfire PM_2.5_ in 20 US states"

### Supplemental Content

**Statistical Analysis Continued**

**eTable 1.** Study periods of each state.

**eTable 2.** The ICD codes used for the disease identification in this study.

**eTable 3.** Sensitivity analysis results adjusting degrees of freedom for temperature to nine.

**eTable 4.** Sensitivity analysis results adjusting for annual precipitation and vapor pressure.

**eTable 5.** Sensitivity analysis results under case definition based on the principal diagnosis code at discharge.

**eTable 6.** Subgroup analysis by age.

**eTable 7.** Subgroup analysis by sex.

**eTable 8.** Subgroup analysis by race/ethnicity.

**eTable 9.** Subgroup analysis by patient location.

**eTable 10.** Subgroup analysis by comorbidity status of obesity.

**eTable 11.** Subgroup analysis by comorbidity status of diabetes.

**eTable 12.** Subgroup analysis by severity of the disease.

**eTable 13.** Subgroup analysis by percentage of the population who graduated high school.

**eTable 14.** Subgroup analysis by townsend deprivation index.

**Statistical Analysis Continued**

In this study, we used conditional logistic regression models to estimate the effects of long-term wildfire-specific and non-wildfire $PM_{2.5}$ on the hospitalization risk, adjusting for measured time-varying confounders, including calendar year, temperature values, and neighborhood-level covariates.

In the conditional logistic regression model, the 2-year average wildfire-specific and non-wildfire $PM_{2.5}$ concentrations were simultaneously included as continuous variables to mutually adjust for each other's effects. Calendar year was included as a categorical variable to control for potential unmeasured confounders varying over time. Surface temperature variables–including mean summer temperature, standard deviation of summer temperature, mean winter temperature, and standard deviation of winter temperature–were modeled using penalized cubic splines with 5 degrees of freedom to account for potential nonlinear relationships. Other covariates included neighborhood-level statistics, such as percentage of White, percentage of Black, percentage of the population who graduated high school, median household income, and Townsend Deprivation Index, all adjusted as continuous variables. The model was provided below.

Let $l(\mu_1(\beta),\cdots,\mu_n(\beta))$ denotes the log likelihood for a conditional logistic regression for observations $i$ = 1,…,n, where

$$\mu_i(\beta) = \beta_1 \boldsymbol{W}_i + \beta_2 \boldsymbol{X}_i + \beta_T \boldsymbol{T}_i + \beta_Y \boldsymbol{Y}_i + \beta_N \boldsymbol{N}_i$$

$\boldsymbol{W}_{i,1}$ and $\boldsymbol{X}_{i,1}$ are the exposure of interest, respectively (2-year average wildfire-specific and non-wildfire $PM_{2.5}$ concentrations, respective), and $\boldsymbol{T}_i$ are the vectors of cubic spline expansions of air temperature. The cubic spline expansion has five degrees of freedom. $\boldsymbol{Y}_i$ and $\boldsymbol{N}_i$ represents calendar year and neighborhood-level statistics, respectively.

**eTable 1.** Study periods of each state.

| State | Abbreviation | Study periods |
|---|---|---|
| Arizona | AZ | 2006-2019 |
| California | CA | 2018-2019 |
| Colorado | CO | 2014-2019 |
| Delaware | DE | 2016-2019 |
| Georgia | GA | 2010-2019 |
| Iowa | IA | 2006-2019 |
| Indiana | IN | 2017-2019 |
| Kentucky | KY | 2006-2019 |
| Maryland | MD | 2006-2019 |
| Michigan | MI | 2006-2019 |
| Minnesota | MN | 2010-2019 |
| North Carolina | NC | 2006-2019 |
| Nebraska | NE | 2014-2019 |
| New Jersey | NJ | 2006-2019 |
| New York | NY | 2006-2019 |
| Oregon | OR | 2006-2019 |
| Rhode Island | RI | 2006-2019 |
| Vermont | VT | 2017-2019 |
| Washington | WA | 2006-2019 |
| Wisconsin | WI | 2012-2019 |

**eTable 2.** The ICD codes used for the disease identification in this study.

| Types of diseases | ICD-9 code | ICD-10 code |
|---|---|---|
| Cardiovascular diseases | | |
| Ischemic heart disease | 410-414 | I20-I25 |
| Cerebrovascular disease | 430-438 | I60-I69 |
| Heart failure | 398.91, 402.01, 402.11, 402.91, 404.01, 404.03, 404.11, 404.13, 404.91, 404.93, 428 | I50 |
| Arrhythmia | 426, 427, 785.0 | I44, I45, I47-I49, R00.0, R00.1, R00.8, R94.31, T82.1, T82.11, T82.110, T82.111, T82.118, T82.119, Z45.0, Z45.01, Z45.010, Z45.018, Z45.02, Z45.09, Z95.0 |
| Hypertension | 401-405 | I10-I15 |
| Other cardiovascular diseases | All other codes within the range 390-459 | All other codes within the range I00-I99 |
| Respiratory diseases | | |
| Acute respiratory infections | 460-466 | J00-J06, J20-J22 |
| Pneumonia | 480-486 | J12-J18 |
| COPD | 491, 492, 496 | J41-J44 |
| Asthma | 493 | J45, J46 |
| Other respiratory diseases | All other codes within the range 460-519 | All other codes within the range J00-J99 |
| Obesity | 2780, 6491 | E66, O99.21 |
| Diabetes | 6480, 249, 250 | E08-E13 |

**eTable 3.** Sensitivity analysis results adjusting degrees of freedom for temperature to nine.

| Outcome | Wildfire-specific $PM_{2.5}$ | Non-wildfire $PM_{2.5}$ |
|---|---|---|
| **CVD** | | |
| Ischemic heart disease | 1.110 (1.100, 1.120) | 1.061 (1.057, 1.064) |
| Cerebrovascular disease | 1.117 (1.105, 1.129) | 1.067 (1.062, 1.071) |
| Heart failure | 1.091 (1.082, 1.099) | 1.053 (1.050, 1.056) |
| Arrhythmia | 1.102 (1.092, 1.112) | 1.049 (1.045, 1.052) |
| Hypertension | 1.111 (1.105, 1.118) | 1.077 (1.075, 1.080) |
| Other cardiovascular diseases | 1.103 (1.095, 1.111) | 1.059 (1.055, 1.062) |
| **RD** | | |
| Acute respiratory infections | 1.098 (1.074, 1.123) | 1.050 (1.041, 1.059) |
| Pneumonia | 1.100 (1.091, 1.109) | 1.054 (1.050, 1.058) |
| COPD | 1.100 (1.089, 1.112) | 1.046 (1.042, 1.050) |
| Asthma | 1.135 (1.118, 1.153) | 1.055 (1.050, 1.061) |
| Other respiratory diseases | 1.108 (1.102, 1.115) | 1.063 (1.060, 1.066) |

The model was adjusted for calendar year, percentage of White, percentage of Black, percentage of the population who graduated high school, median household income, townsend deprivation index, mean temperature of summer and winter, and standard deviation of summer and winter temperature.

**eTable 4.** Sensitivity analysis results adjusting for annual precipitation and vapor pressure.

| Outcome | Wildfire-specific $PM_{2.5}$ | Non-wildfire $PM_{2.5}$ |
|---|---|---|
| **CVD** | | |
| Ischemic heart disease | 1.105 (1.095, 1.115) | 1.062 (1.058, 1.066) |
| Cerebrovascular disease | 1.112 (1.100, 1.124) | 1.069 (1.065, 1.074) |
| Heart failure | 1.083 (1.074, 1.092) | 1.054 (1.051, 1.057) |
| Arrhythmia | 1.100 (1.090, 1.110) | 1.050 (1.047, 1.054) |
| Hypertension | 1.096 (1.090, 1.103) | 1.082 (1.079, 1.085) |
| Other cardiovascular diseases | 1.102 (1.094, 1.111) | 1.061 (1.058, 1.064) |
| **RD** | | |
| Acute respiratory infections | 1.103 (1.078, 1.128) | 1.052 (1.044, 1.061) |
| Pneumonia | 1.096 (1.087, 1.106) | 1.055 (1.052, 1.059) |
| COPD | 1.093 (1.081, 1.104) | 1.045 (1.040, 1.049) |
| Asthma | 1.134 (1.116, 1.152) | 1.056 (1.051, 1.062) |
| Other respiratory diseases | 1.101 (1.094, 1.108) | 1.066 (1.063, 1.069) |

The model was adjusted for calendar year, percentage of White, percentage of Black, percentage of the population who graduated high school, median household income, townsend deprivation index, mean temperature of summer and winter, standard deviation of summer and winter temperature, annual precipitation, and annual vapor pressure. Daily precipitation vapor pressure (Pa) were derived from Daymet V4.

**eTable 5.** Sensitivity analysis results under case definition based on the principal diagnosis code at discharge.

| Outcome | Wildfire-specific $PM_{2.5}$ | Non-wildfire $PM_{2.5}$ |
|---|---|---|
| **CVD** | | |
| Ischemic heart disease | 1.150 (1.136, 1.164) | 1.071 (1.066, 1.076) |
| Cerebrovascular disease | 1.137 (1.124, 1.151) | 1.079 (1.073, 1.084) |
| Heart failure | 1.092 (1.072, 1.113) | 1.020 (1.014, 1.027) |
| Arrhythmia | 1.118 (1.102, 1.135) | 1.057 (1.051, 1.062) |
| Hypertension | 1.278 (1.259, 1.297) | 1.148 (1.140, 1.156) |
| Other cardiovascular diseases | 1.144 (1.130, 1.157) | 1.069 (1.064, 1.074) |
| **RD** | | |
| Acute respiratory infections | 1.125 (1.094, 1.158) | 1.052 (1.041, 1.063) |
| Pneumonia | 1.086 (1.070, 1.101) | 1.049 (1.044, 1.055) |
| COPD | 1.141 (1.122, 1.160) | 1.060 (1.053, 1.067) |
| Asthma | 1.158 (1.126, 1.191) | 1.056 (1.046, 1.065) |
| Other respiratory diseases | 1.117 (1.104, 1.130) | 1.071 (1.066, 1.076) |

The model was adjusted for calendar year, percentage of White, percentage of Black, percentage of the population who graduated high school, median household income, townsend deprivation index, mean temperature of summer and winter, and standard deviation of summer and winter temperature.

**eTable 6.** Subgroup analysis by age.

| Outcome | Wildfire-specific $PM_{2.5}$ | | | | Non-wildfire $PM_{2.5}$ | | |
|---|---|---|---|---|---|---|---|
| | Under 65 years old | 65 years old and above | *P*-value[a] | | Under 65 years old | 65 years old and above | *P*-value[b] |
| **CVD** | | | | | | | |
| Ischemic heart disease | 1.129 (1.112, 1.146) | 1.130 (1.118, 1.142) | 0.876 | | 1.067 (1.061, 1.072) | 1.064 (1.059, 1.068) | 0.247 |
| Cerebrovascular disease | 1.141 (1.120, 1.162) | 1.127 (1.113, 1.142) | 0.142 | | 1.081 (1.073, 1.089) | 1.067 (1.061, 1.072) | <0.001 |
| Heart failure | 1.144 (1.127, 1.162) | 1.086 (1.076, 1.096) | <0.001 | | 1.079 (1.072, 1.085) | 1.046 (1.042, 1.050) | <0.001 |
| Arrhythmia | 1.141 (1.121, 1.162) | 1.110 (1.099, 1.121) | <0.001 | | 1.061 (1.054, 1.068) | 1.049 (1.044, 1.053) | <0.001 |
| Hypertension | 1.121 (1.110, 1.131) | 1.130 (1.121, 1.139) | 0.054 | | 1.084 (1.081, 1.088) | 1.085 (1.081, 1.088) | 0.907 |
| Other cardiovascular diseases | 1.127 (1.115, 1.140) | 1.111 (1.100, 1.122) | 0.005 | | 1.072 (1.067, 1.076) | 1.056 (1.052, 1.060) | <0.001 |
| **RD** | | | | | | | |
| Acute respiratory infections [c] | 1.123 (1.092, 1.156) | 1.089 (1.052, 1.126) | 0.045 | | 1.050 (1.039, 1.061) | 1.062 (1.049, 1.075) | 0.053 |
| Pneumonia | 1.105 (1.090, 1.120) | 1.111 (1.099, 1.123) | 0.356 | | 1.056 (1.051, 1.062) | 1.056 (1.052, 1.061) | 0.980 |
| COPD | 1.103 (1.083, 1.122) | 1.119 (1.105, 1.132) | 0.057 | | 1.047 (1.040, 1.054) | 1.047 (1.042, 1.052) | 0.948 |
| Asthma [c] | 1.165 (1.127, 1.205) | 1.158 (1.138, 1.178) | 0.645 | | 1.067 (1.055, 1.079) | 1.058 (1.051, 1.064) | 0.052 |
| Other respiratory diseases | 1.116 (1.106, 1.126) | 1.113 (1.104, 1.121) | 0.474 | | 1.071 (1.067, 1.075) | 1.062 (1.059, 1.066) | <0.001 |

The model was adjusted for calendar year, percentage of White, percentage of Black, percentage of the population who graduated high school, median household income, townsend deprivation index, mean temperature of summer and winter, standard deviation of summer and winter.

[a] The comparison of wildfire-specific $PM_{2.5}$ effect values between subgroups. *P*-value was calculated by Z-test.

[b] The comparison of non-wildfire $PM_{2.5}$ effect values between subgroups. *P*-value was calculated by Z-test.

[c] The group was divided into under 18 years old vs. 18 years old and above.

**eTable 7.** Subgroup analysis by sex.

| Outcome | Wildfire-specific $PM_{2.5}$ | | | | Non-wildfire $PM_{2.5}$ | | |
|---|---|---|---|---|---|---|---|
| | Female | Male | *P*-value[a] | | Female | Male | *P*-value[b] |
| **CVD** | | | | | | | |
| Ischemic heart disease | 1.132 (1.116, 1.148) | 1.127 (1.114, 1.139) | 0.441 | | 1.067 (1.062, 1.073) | 1.063 (1.059, 1.068) | 0.134 |
| Cerebrovascular disease | 1.131 (1.115, 1.148) | 1.130 (1.114, 1.147) | 0.868 | | 1.070 (1.064, 1.076) | 1.072 (1.066, 1.079) | 0.422 |
| Heart failure | 1.093 (1.080, 1.105) | 1.106 (1.094, 1.118) | 0.032 | | 1.052 (1.047, 1.057) | 1.057 (1.053, 1.062) | 0.024 |
| Arrhythmia | 1.122 (1.108, 1.137) | 1.114 (1.100, 1.127) | 0.223 | | 1.053 (1.047, 1.058) | 1.051 (1.046, 1.056) | 0.453 |
| Hypertension | 1.121 (1.111, 1.131) | 1.124 (1.114, 1.134) | 0.510 | | 1.082 (1.078, 1.086) | 1.088 (1.084, 1.092) | 0.002 |
| Other cardiovascular diseases | 1.116 (1.105, 1.128) | 1.119 (1.107, 1.130) | 0.727 | | 1.062 (1.057, 1.066) | 1.063 (1.059, 1.067) | 0.530 |
| **RD** | | | | | | | |
| Acute respiratory infections | 1.101 (1.068, 1.136) | 1.119 (1.085, 1.154) | 0.305 | | 1.053 (1.041, 1.066) | 1.054 (1.042, 1.066) | 0.914 |
| Pneumonia | 1.108 (1.095, 1.121) | 1.109 (1.096, 1.123) | 0.822 | | 1.054 (1.049, 1.059) | 1.058 (1.053, 1.063) | 0.138 |
| COPD | 1.117 (1.101, 1.132) | 1.107 (1.091, 1.124) | 0.235 | | 1.046 (1.041, 1.052) | 1.047 (1.041, 1.053) | 0.811 |
| Asthma | 1.156 (1.134, 1.178) | 1.165 (1.135, 1.197) | 0.479 | | 1.060 (1.053, 1.067) | 1.059 (1.049, 1.068) | 0.748 |
| Other respiratory diseases | 1.115 (1.106, 1.125) | 1.113 (1.104, 1.122) | 0.579 | | 1.064 (1.061, 1.068) | 1.068 (1.064, 1.071) | 0.101 |

The model was adjusted for calendar year, percentage of White, percentage of Black, percentage of the population who graduated high school, median household income, townsend deprivation index, mean temperature of summer and winter, standard deviation of summer and winter.

[a] The comparison of wildfire-specific $PM_{2.5}$ effect values between subgroups. *P*-value was calculated by Z-test.

[b] The comparison of non-wildfire $PM_{2.5}$ effect values between subgroups. *P*-value was calculated by Z-test.

**eTable 8.** Subgroup analysis by race/ethnicity.

| Outcome | Wildfire-specific $PM_{2.5}$ | | | | | Non-wildfire $PM_{2.5}$ | | | | |
|---|---|---|---|---|---|---|---|---|---|---|
| | White | Black | Hispanic | Other | *P*-value | White | Black | Hispanic | Other | *P*-value |
| **CVD** | | | | | | | | | | |
| Ischemic heart disease | 1.095 (1.084, 1.106) | 1.061 (1.017, 1.106) | 1.199 (1.150, 1.249) | 1.309 (1.258, 1.363) | <0.001 | 1.070 (1.066, 1.075) | 1.112 (1.098, 1.126) | 1.107 (1.091, 1.124) | 1.137 (1.122, 1.153) | <0.001 |
| Cerebrovascular disease | 1.106 (1.093, 1.120) | 1.077 (1.034, 1.123) | 1.138 (1.089, 1.190) | 1.289 (1.232, 1.349) | <0.001 | 1.073 (1.067, 1.079) | 1.117 (1.102, 1.132) | 1.105 (1.085, 1.125) | 1.165 (1.145, 1.185) | <0.001 |
| Heart failure | 1.073 (1.063, 1.082) | 1.065 (1.033, 1.097) | 1.123 (1.080, 1.167) | 1.169 (1.125, 1.216) | <0.001 | 1.053 (1.049, 1.057) | 1.100 (1.089, 1.110) | 1.063 (1.048, 1.079) | 1.128 (1.112, 1.145) | <0.001 |
| Arrhythmia | 1.098 (1.087, 1.109) | 1.090 (1.040, 1.143) | 1.156 (1.101, 1.213) | 1.212 (1.156, 1.272) | <0.001 | 1.063 (1.059, 1.068) | 1.092 (1.077, 1.108) | 1.079 (1.060, 1.098) | 1.119 (1.101, 1.138) | <0.001 |
| Hypertension | 1.076 (1.068, 1.084) | 1.059 (1.036, 1.083) | 1.195 (1.165, 1.227) | 1.308 (1.271, 1.346) | <0.001 | 1.079 (1.076, 1.083) | 1.119 (1.112, 1.127) | 1.102 (1.091, 1.112) | 1.153 (1.141, 1.165) | <0.001 |
| Other cardiovascular diseases | 1.093 (1.084, 1.103) | 1.075 (1.043, 1.107) | 1.160 (1.123, 1.199) | 1.238 (1.195, 1.283) | <0.001 | 1.064 (1.061, 1.068) | 1.102 (1.092, 1.112) | 1.105 (1.091, 1.119) | 1.136 (1.121, 1.151) | <0.001 |
| **RD** | | | | | | | | | | |
| Acute respiratory infections | 1.061 (1.031, 1.092) | 1.100 (1.014, 1.193) | 1.173 (1.108, 1.241) | 1.253 (1.157, 1.356) | <0.001 | 1.046 (1.034, 1.058) | 1.109 (1.083, 1.137) | 1.086 (1.064, 1.110) | 1.109 (1.078, 1.140) | <0.001 |
| Pneumonia | 1.088 (1.077, 1.098) | 1.062 (1.022, 1.103) | 1.149 (1.108, 1.191) | 1.237 (1.191, 1.285) | <0.001 | 1.054 (1.049, 1.058) | 1.106 (1.093, 1.119) | 1.094 (1.079, 1.109) | 1.133 (1.117, 1.150) | <0.001 |
| COPD | 1.087 (1.075, 1.099) | 1.102 (1.050, 1.157) | 1.190 (1.115, 1.270) | 1.240 (1.171, 1.314) | <0.001 | 1.042 (1.037, 1.047) | 1.125 (1.108, 1.142) | 1.066 (1.042, 1.091) | 1.121 (1.098, 1.146) | <0.001 |
| Asthma | 1.120 (1.098, 1.143) | 1.143 (1.091, 1.199) | 1.243 (1.184, 1.304) | 1.280 (1.204, 1.361) | <0.001 | 1.059 (1.051, 1.067) | 1.088 (1.074, 1.102) | 1.084 (1.067, 1.101) | 1.108 (1.086, 1.130) | <0.001 |
| Other respiratory diseases | 1.089 (1.082, 1.096) | 1.089 (1.061, 1.118) | 1.143 (1.115, 1.173) | 1.264 (1.230, 1.298) | <0.001 | 1.066 (1.062, 1.069) | 1.119 (1.109, 1.129) | 1.108 (1.096, 1.119) | 1.154 (1.141, 1.166) | <0.001 |

The model was adjusted for calendar year, percentage of White, percentage of Black, percentage of the population who graduated high school, median household income, mean temperature of summer and winter, standard deviation of summer and winter.

[a] The comparison of wildfire-specific $PM_{2.5}$ effect values between subgroups. *P*-value was calculated by Wald test.

[b] The comparison of non-wildfire $PM_{2.5}$ effect values between subgroups. *P*-value was calculated by Wald test.

**eTable 9.** Subgroup analysis by patient location.

| Outcome | Wildfire-specific $PM_{2.5}$ | | | | Non-wildfire $PM_{2.5}$ | | | |
|---|---|---|---|---|---|---|---|---|
| | Metropolitan | Micropolitan | Non-CBSA | *P*-value[a] | Metropolitan | Micropolitan | Non-CBSA | *P*-value[b] |
| **CVD** | | | | | | | | |
| Ischemic heart disease | 1.144 (1.133, 1.156) | 1.034 (1.008, 1.060) | 1.017 (0.986, 1.050) | <0.001 | 1.069 (1.065, 1.073) | 1.014 (1.004, 1.024) | 0.994 (0.981, 1.007) | <0.001 |
| Cerebrovascular disease | 1.146 (1.133, 1.160) | 1.046 (1.014, 1.079) | 1.017 (0.978, 1.057) | <0.001 | 1.074 (1.069, 1.079) | 1.006 (0.992, 1.020) | 1.015 (0.996, 1.033) | <0.001 |
| Heart failure | 1.106 (1.097, 1.116) | 1.029 (1.006, 1.052) | 0.993 (0.963, 1.023) | <0.001 | 1.054 (1.050, 1.057) | 1.010 (1.000, 1.020) | 1.007 (0.994, 1.020) | <0.001 |
| Arrhythmia | 1.129 (1.118, 1.140) | 1.032 (1.006, 1.059) | 1.033 (1.000, 1.067) | <0.001 | 1.054 (1.050, 1.058) | 1.001 (0.990, 1.012) | 1.005 (0.991, 1.019) | <0.001 |
| Hypertension | 1.131 (1.123, 1.138) | 1.034 (1.016, 1.053) | 1.031 (1.007, 1.056) | <0.001 | 1.089 (1.086, 1.092) | 1.002 (0.994, 1.011) | 1.002 (0.991, 1.013) | <0.001 |
| Other cardiovascular diseases | 1.128 (1.119, 1.138) | 1.043 (1.021, 1.066) | 1.026 (0.998, 1.055) | <0.001 | 1.064 (1.061, 1.067) | 1.006 (0.996, 1.016) | 1.007 (0.995, 1.021) | <0.001 |
| **RD** | | | | | | | | |
| Acute respiratory infections | 1.121 (1.094, 1.149) | 1.061 (0.992, 1.134) | 1.040 (0.954, 1.133) | 0.008 | 1.056 (1.047, 1.065) | 0.988 (0.961, 1.016) | 0.976 (0.942, 1.012) | <0.001 |
| Pneumonia | 1.118 (1.108, 1.129) | 1.052 (1.027, 1.077) | 1.015 (0.985, 1.045) | <0.001 | 1.059 (1.055, 1.063) | 0.995 (0.985, 1.005) | 0.991 (0.978, 1.004) | <0.001 |
| COPD | 1.123 (1.110, 1.137) | 1.047 (1.019, 1.075) | 1.050 (1.015, 1.086) | <0.001 | 1.054 (1.049, 1.058) | 1.005 (0.993, 1.016) | 0.996 (0.982, 1.010) | <0.001 |
| Asthma | 1.175 (1.155, 1.195) | 1.041 (0.992, 1.092) | 1.026 (0.959, 1.098) | <0.001 | 1.060 (1.054, 1.066) | 1.023 (1.003, 1.043) | 1.010 (0.983, 1.037) | <0.001 |
| Other respiratory diseases | 1.120 (1.113, 1.127) | 1.074 (1.056, 1.092) | 1.040 (1.016, 1.065) | <0.001 | 1.069 (1.066, 1.072) | 0.996 (0.988, 1.005) | 0.999 (0.988, 1.010) | <0.001 |

The model was adjusted for calendar year, percentage of White, percentage of Black, percentage of the population who graduated high school, median household income, mean temperature of summer and winter, standard deviation of summer and winter.

[a] The comparison of wildfire-specific $PM_{2.5}$ effect values between subgroups. *P*-value was calculated by Wald test.

[b] The comparison of non-wildfire $PM_{2.5}$ effect values between subgroups. *P*-value was calculated by Wald test.

**eTable 10.** Subgroup analysis by ICD-9-CM diagnoses of obesity.

| Outcome | Wildfire-specific $PM_{2.5}$ | | | | Non-wildfire $PM_{2.5}$ | | |
|---|---|---|---|---|---|---|---|
| | Obesity | Non-obesity | *P*-value[a] | | Obesity | Non-obesity | *P*-value[b] |
| **CVD** | | | | | | | |
| Ischemic heart disease | 1.184 (1.152, 1.216) | 1.123 (1.112, 1.133) | <0.001 | | 1.081 (1.069, 1.093) | 1.063 (1.059, 1.067) | <0.001 |
| Cerebrovascular disease | 1.196 (1.148, 1.247) | 1.127 (1.115, 1.139) | <0.001 | | 1.101 (1.082, 1.120) | 1.069 (1.065, 1.074) | <0.001 |
| Heart failure | 1.121 (1.095, 1.148) | 1.097 (1.088, 1.106) | 0.013 | | 1.074 (1.063, 1.085) | 1.053 (1.049, 1.056) | <0.001 |
| Arrhythmia | 1.161 (1.129, 1.195) | 1.113 (1.102, 1.123) | <0.001 | | 1.061 (1.049, 1.074) | 1.051 (1.047, 1.055) | 0.026 |
| Hypertension | 1.102 (1.082, 1.121) | 1.125 (1.117, 1.132) | 0.001 | | 1.090 (1.082, 1.098) | 1.084 (1.081, 1.087) | 0.046 |
| Other cardiovascular diseases | 1.150 (1.123, 1.177) | 1.114 (1.105, 1.123) | <0.001 | | 1.073 (1.062, 1.084) | 1.061 (1.058, 1.065) | 0.003 |
| **RD** | | | | | | | |
| Acute respiratory infections | 1.125 (1.016, 1.246) | 1.110 (1.086, 1.135) | 0.720 | | 1.071 (1.031, 1.113) | 1.053 (1.044, 1.062) | 0.222 |
| Pneumonia | 1.146 (1.107, 1.185) | 1.106 (1.097, 1.116) | 0.005 | | 1.068 (1.054, 1.083) | 1.055 (1.052, 1.059) | 0.014 |
| COPD | 1.151 (1.115, 1.189) | 1.108 (1.096, 1.120) | 0.001 | | 1.065 (1.051, 1.079) | 1.045 (1.040, 1.049) | <0.001 |
| Asthma | 1.158 (1.108, 1.211) | 1.159 (1.140, 1.179) | 0.950 | | 1.059 (1.043, 1.076) | 1.060 (1.054, 1.066) | 0.952 |
| Other respiratory diseases | 1.147 (1.124, 1.170) | 1.111 (1.104, 1.118) | <0.001 | | 1.074 (1.064, 1.084) | 1.065 (1.062, 1.068) | 0.020 |

The model was adjusted for calendar year, percentage of White, percentage of Black, percentage of the population who graduated high school, median household income, townsend deprivation index, mean temperature of summer and winter, standard deviation of summer and winter.

[a] The comparison of wildfire-specific $PM_{2.5}$ effect values between subgroups. *P*-value was calculated by Z-test.

[b] The comparison of non-wildfire $PM_{2.5}$ effect values between subgroups. *P*-value was calculated by Z-test.

**eTable 11.** Subgroup analysis by comorbidity status of diabetes.

| Outcome | Wildfire-specific $PM_{2.5}$ | | | | Non-wildfire $PM_{2.5}$ | | |
|---|---|---|---|---|---|---|---|
| | Diabetes | Non-diabetes | *P*-value[a] | | Diabetes | Non-diabetes | *P*-value[b] |
| **CVD** | | | | | | | |
| Ischemic heart disease | 1.144 (1.125, 1.162) | 1.122 (1.110, 1.134) | 0.004 | | 1.073 (1.067, 1.080) | 1.061 (1.057, 1.065) | <0.001 |
| Cerebrovascular disease | 1.151 (1.128, 1.175) | 1.123 (1.110, 1.137) | 0.003 | | 1.085 (1.076, 1.094) | 1.066 (1.061, 1.071) | <0.001 |
| Heart failure | 1.106 (1.091, 1.121) | 1.096 (1.085, 1.106) | 0.109 | | 1.059 (1.054, 1.065) | 1.052 (1.048, 1.056) | 0.004 |
| Arrhythmia | 1.114 (1.093, 1.135) | 1.118 (1.107, 1.129) | 0.582 | | 1.053 (1.045, 1.061) | 1.051 (1.047, 1.055) | 0.498 |
| Hypertension | 1.116 (1.103, 1.129) | 1.119 (1.111, 1.127) | 0.600 | | 1.100 (1.094, 1.105) | 1.078 (1.075, 1.081) | 0 |
| Other cardiovascular diseases | 1.110 (1.092, 1.127) | 1.119 (1.110, 1.128) | 0.178 | | 1.062 (1.055, 1.068) | 1.062 (1.059, 1.066) | 0.776 |
| **RD** | | | | | | | |
| Acute respiratory infections | 1.068 (0.993, 1.150) | 1.114 (1.089, 1.140) | 0.120 | | 1.059 (1.031, 1.089) | 1.054 (1.045, 1.062) | 0.595 |
| Pneumonia | 1.123 (1.103, 1.144) | 1.104 (1.094, 1.115) | 0.020 | | 1.066 (1.058, 1.073) | 1.053 (1.049, 1.057) | <0.001 |
| COPD | 1.111 (1.087, 1.134) | 1.113 (1.100, 1.126) | 0.821 | | 1.048 (1.040, 1.057) | 1.046 (1.041, 1.051) | 0.437 |
| Asthma | 1.147 (1.095, 1.202) | 1.161 (1.142, 1.180) | 0.505 | | 1.039 (1.024, 1.055) | 1.062 (1.056, 1.069) | <0.001 |
| Other respiratory diseases | 1.134 (1.119, 1.148) | 1.109 (1.102, 1.116) | <0.001 | | 1.073 (1.067, 1.079) | 1.064 (1.061, 1.067) | <0.001 |

The model was adjusted for calendar year, percentage of White, percentage of Black, percentage of the population who graduated high school, median household income, townsend deprivation index, mean temperature of summer and winter, standard deviation of summer and winter.

[a] The comparison of wildfire-specific $PM_{2.5}$ effect values between subgroups. *P*-value was calculated by Z-test.

[b] The comparison of non-wildfire $PM_{2.5}$ effect values between subgroups. *P*-value was calculated by Z-test.

**eTable 12.** Subgroup analysis by severity of the disease.

| Outcome | Wildfire-specific $PM_{2.5}$ | | | | Non-wildfire $PM_{2.5}$ | | |
|---|---|---|---|---|---|---|---|
| | Died during hospitalization | Not died during hospitalization | *P*-value[a] | | Died during hospitalization | Not died during hospitalization | *P*-value[b] |
| **CVD** | | | | | | | |
| Ischemic heart disease | 1.096 (1.051, 1.143) | 1.130 (1.120, 1.140) | 0.045 | | 1.080 (1.061, 1.099) | 1.064 (1.060, 1.068) | 0.016 |
| Cerebrovascular disease | 1.158 (1.108, 1.210) | 1.129 (1.117, 1.141) | 0.115 | | 1.082 (1.062, 1.102) | 1.070 (1.066, 1.075) | 0.113 |
| Heart failure | 1.112 (1.064, 1.162) | 1.099 (1.090, 1.107) | 0.445 | | 1.054 (1.035, 1.074) | 1.054 (1.051, 1.058) | 0.990 |
| Arrhythmia | 1.099 (1.033, 1.168) | 1.117 (1.107, 1.127) | 0.443 | | 1.047 (1.023, 1.071) | 1.051 (1.048, 1.055) | 0.618 |
| Hypertension | 1.254 (1.175, 1.339) | 1.120 (1.114, 1.127) | <0.001 | | 1.114 (1.081, 1.149) | 1.084 (1.082, 1.087) | 0.011 |
| Other cardiovascular diseases | 1.110 (1.065, 1.157) | 1.117 (1.109, 1.126) | 0.662 | | 1.061 (1.042, 1.080) | 1.062 (1.059, 1.065) | 0.856 |
| **RD** | | | | | | | |
| Acute respiratory infections | 1.301 (0.703, 2.407) | 1.110 (1.086, 1.134) | 0.463 | | 1.148 (0.892, 1.477) | 1.053 (1.045, 1.062) | 0.335 |
| Pneumonia | 1.094 (1.053, 1.137) | 1.109 (1.099, 1.119) | 0.344 | | 1.062 (1.046, 1.078) | 1.055 (1.052, 1.059) | 0.252 |
| COPD | 1.087 (0.996, 1.185) | 1.112 (1.101, 1.124) | 0.449 | | 1.038 (1.004, 1.073) | 1.046 (1.042, 1.050) | 0.487 |
| Asthma | 1.098 (0.788, 1.529) | 1.159 (1.142, 1.177) | 0.643 | | 1.003 (0.896, 1.122) | 1.060 (1.054, 1.065) | 0.164 |
| Other respiratory diseases | 1.120 (1.099, 1.142) | 1.113 (1.106, 1.120) | 0.367 | | 1.075 (1.066, 1.084) | 1.065 (1.062, 1.068) | 0.002 |

The model was adjusted for calendar year, percentage of White, percentage of Black, percentage of the population who graduated high school, median household income, townsend deprivation index, mean temperature of summer and winter, standard deviation of summer and winter.

[a] The comparison of wildfire-specific $PM_{2.5}$ effect values between subgroups. *P*-value was calculated by Z-test.

[b] The comparison of non-wildfire $PM_{2.5}$ effect values between subgroups. *P*-value was calculated by Z-test.

**eTable 13.** Subgroup analysis by percentage of the population who graduated high school.

| Outcome | Wildfire-specific $PM_{2.5}$ | | | | Non-wildfire $PM_{2.5}$ | | |
|---|---|---|---|---|---|---|---|
| | Percentage of the population who graduated high school ≤ 25th | Percentage of the population who graduated high school > 25th | $P$-value[a] | | Percentage of the population who graduated high school ≤ 25th | Percentage of the population who graduated high school > 25th | $P$-value[b] |
| **CVD** | | | | | | | |
| Ischemic heart disease | 1.181 (1.154, 1.209) | 1.126 (1.115, 1.137) | <0.001 | | 1.107 (1.099, 1.115) | 1.050 (1.046, 1.054) | <0.001 |
| Cerebrovascular disease | 1.205 (1.172, 1.238) | 1.125 (1.113, 1.138) | <0.001 | | 1.125 (1.115, 1.136) | 1.052 (1.047, 1.057) | <0.001 |
| Heart failure | 1.179 (1.154, 1.204) | 1.092 (1.083, 1.102) | <0.001 | | 1.098 (1.090, 1.106) | 1.036 (1.032, 1.040) | <0.001 |
| Arrhythmia | 1.187 (1.161, 1.214) | 1.118 (1.107, 1.129) | <0.001 | | 1.096 (1.088, 1.104) | 1.037 (1.033, 1.041) | <0.001 |
| Hypertension | 1.245 (1.225, 1.266) | 1.112 (1.105, 1.120) | <0.001 | | 1.130 (1.124, 1.136) | 1.062 (1.059, 1.066) | <0.001 |
| Other cardiovascular diseases | 1.169 (1.147, 1.192) | 1.114 (1.105, 1.123) | <0.001 | | 1.119 (1.112, 1.126) | 1.044 (1.040, 1.047) | <0.001 |
| **RD** | | | | | | | |
| Acute respiratory infections | 1.125 (1.064, 1.189) | 1.111 (1.084, 1.138) | 0.561 | | 1.072 (1.054, 1.091) | 1.043 (1.033, 1.053) | <0.001 |
| Pneumonia | 1.159 (1.134, 1.185) | 1.110 (1.100, 1.121) | <0.001 | | 1.095 (1.087, 1.103) | 1.040 (1.036, 1.044) | <0.001 |
| COPD | 1.193 (1.162, 1.225) | 1.108 (1.096, 1.121) | <0.001 | | 1.087 (1.078, 1.097) | 1.033 (1.028, 1.037) | <0.001 |
| Asthma | 1.213 (1.163, 1.265) | 1.160 (1.140, 1.179) | 0.005 | | 1.094 (1.081, 1.108) | 1.049 (1.043, 1.056) | <0.001 |
| Other respiratory diseases | 1.192 (1.175, 1.211) | 1.105 (1.098, 1.112) | <0.001 | | 1.124 (1.118, 1.130) | 1.044 (1.040, 1.047) | <0.001 |

The model was adjusted for calendar year, percentage of White, percentage of Black, median household income, townsend deprivation index, mean temperature of summer and winter, standard deviation of summer and winter.

[a] The comparison of wildfire-specific $PM_{2.5}$ effect values between subgroups. $P$-value was calculated by Z-test.

[b] The comparison of non-wildfire $PM_{2.5}$ effect values between subgroups. $P$-value was calculated by Z-test.

**eTable 14.** Subgroup analysis by townsend deprivation index.

| Outcome | Wildfire-specific $PM_{2.5}$ | | | Non-wildfire $PM_{2.5}$ | | |
|---|---|---|---|---|---|---|
| | Townsend deprivation index ≤ 75th | Townsend deprivation index > 75th | *P*-value[a] | Townsend deprivation index ≤ 75th | Townsend deprivation index > 75th | *P*-value[b] |
| **CVD** | | | | | | |
| Ischemic heart disease | 1.104 (1.093, 1.115) | 1.226 (1.199, 1.253) | <0.001 | 1.042 (1.038, 1.046) | 1.108 (1.101, 1.116) | <0.001 |
| Cerebrovascular disease | 1.105 (1.092, 1.118) | 1.219 (1.187, 1.252) | <0.001 | 1.047 (1.042, 1.052) | 1.123 (1.114, 1.133) | <0.001 |
| Heart failure | 1.078 (1.069, 1.088) | 1.180 (1.155, 1.205) | <0.001 | 1.034 (1.030, 1.038) | 1.100 (1.093, 1.107) | <0.001 |
| Arrhythmia | 1.085 (1.074, 1.096) | 1.235 (1.209, 1.261) | <0.001 | 1.031 (1.027, 1.035) | 1.097 (1.089, 1.104) | <0.001 |
| Hypertension | 1.103 (1.096, 1.110) | 1.240 (1.217, 1.264) | <0.001 | 1.061 (1.058, 1.064) | 1.137 (1.131, 1.143) | <0.001 |
| Other cardiovascular diseases | 1.090 (1.082, 1.099) | 1.229 (1.206, 1.252) | <0.001 | 1.039 (1.035, 1.042) | 1.117 (1.110, 1.124) | <0.001 |
| **RD** | | | | | | |
| Acute respiratory infections | 1.089 (1.064, 1.115) | 1.223 (1.141, 1.310) | <0.001 | 1.040 (1.030, 1.049) | 1.072 (1.053, 1.092) | <0.001 |
| Pneumonia | 1.078 (1.068, 1.088) | 1.198 (1.173, 1.224) | <0.001 | 1.030 (1.026, 1.034) | 1.103 (1.096, 1.111) | <0.001 |
| COPD | 1.080 (1.068, 1.093) | 1.210 (1.181, 1.239) | <0.001 | 1.020 (1.015, 1.024) | 1.103 (1.094, 1.112) | <0.001 |
| Asthma | 1.150 (1.131, 1.169) | 1.163 (1.088, 1.243) | 0.635 | 1.059 (1.053, 1.065) | 1.055 (1.042, 1.069) | 0.481 |
| Other respiratory diseases | 1.090 (1.083, 1.097) | 1.214 (1.197, 1.231) | <0.001 | 1.035 (1.032, 1.038) | 1.131 (1.126, 1.137) | <0.001 |

The model was adjusted for calendar year, percentage of White, percentage of Black, percentage of the population who graduated high school, median household income, mean temperature of summer and winter, standard deviation of summer and winter.

[a] The comparison of wildfire-specific $PM_{2.5}$ effect values between subgroups. *P*-value was calculated by Z-test.

[b] The comparison of non-wildfire $PM_{2.5}$ effect values between subgroups. *P*-value was calculated by Z-test.
